# Soluble angiotensin-converting enzyme 2 is transiently elevated in COVID-19 and correlates with specific inflammatory and endothelial markers

**DOI:** 10.1101/2021.03.03.21252841

**Authors:** Annika Lundström, Louise Ziegler, Sebastian Havervall, Ann-Sofie Rudberg, Fien von Meijenfeldt, Ton Lisman, Nigel Mackman, Per Sandén, Charlotte Thålin

## Abstract

**Rationale:** Angiotensin-converting enzyme 2 (ACE2) is the main entry receptor of severe acute respiratory syndrome coronavirus 2 (SARS-CoV-2), but how SARS-CoV-2 interactions with ACE2 influences the renin-angiotensin system (RAS) in Coronavirus disease 2019 (COVID-19) is unknown.

**Objective:** To measure circulating ACE2 and ACE levels in COVID-19 patients and investigate association with risk factors, outcome and inflammatory markers.

**Methods and results:** Soluble ACE2 (sACE2) and sACE concentrations were measured by ELISA in plasma samples from 114 hospital-treated COVID-19 patients and 10 healthy controls. Follow-up samples after four months were available for 58/114 patients. Von Willebrand factor (VWF), factor VIII (fVIII), D-dimer, interleukin 6 (IL-6), tumor necrosis factor α and plasminogen activator inhibitor 1 (PAI-1) had previously been determined. Levels of sACE2 were higher in COVID-19 patients than in healthy controls, median 5.0 (interquartile range 2.8-11.8) ng/ml versus 1.4 (1.1-1.6) ng/ml, p < 0.0001. sACE2 was higher in men than women, but were not affected by other risk factors for severe COVID-19. sACE 2 decreased to 2.3 (1.6-3.9) ng/ml at follow-up, p < 0.0001, but remained higher than in healthy controls, p=0.012. Follow-up sACE2 levels were higher with increasing age, BMI, total number of comorbidities, for patients with diabetes and patients on RAS-inhibition. sACE was marginally lower during COVID-19 compared with at follow-up, 57 (45-70) ng/ml versus 72 (52-87) ng/ml, p=0.008. Levels of sACE2 and sACE did not differ depending on survival or disease severity (care level, respiratory support). sACE2 during COVID-19 correlated with VWF, fVIII and D-dimer, while sACE correlated with IL-6, TNFα and PAI-1.

**Conclusions:** sACE2 was transiently elevated in COVID-19, likely due to increased shedding from infected cells. sACE2 and sACE during COVID-19 differed distinctly in their correlations with markers of inflammation and endothelial dysfunction, suggesting release from different cell types and/or vascular beds.

## 1 Introduction

Angiotensin-converting enzyme (ACE) 2, ACE2, is the main entry receptor for severe acute respiratory syndrome coronavirus 2 (SARS-CoV-2), the virus causing the ongoing pandemic with at the time of writing more than 110 million infected and 2.4 million dead worldwide. ACE2 is a homologue of ACE and was discovered by two independent groups in 2000 (1, 2). Importantly, ACE2 cleaves angiotensin II (Ang II) into angiotensin 1-7 (Ang 1-7), counteracting Ang II in the classical renin-angiotensin system (RAS) (3, 4). The complete RAS system include several enzymes, angiotensin peptides and receptors, with complex interactions and feed-forward/feedback mechanisms (3). The main RAS effector arms are considered to be ACE/Ang II/angiotensin 1 receptor (AT1R), mediating vasoconstrictive, inflammatory, proliferative and pro-thrombotic effects, and ACE2/Ang 1-7/Mas receptor (MasR), which has opposite effects (5-7), see figure 1. ACE2 and ACE are both membrane-anchored enzymes with wide tissue distribution (8, 9). The presence of ACE2 on different cells is thought to contribute to the varying organ manifestations seen in Coronavirus disease 2019 (COVID-19) (3, 10). Both ACE2 and ACE can be shed from the cell surface to soluble forms that retain enzyme activity (6, 11). In addition to systemic RAS, which is central for cardiovascular regulation, many organs including the lungs have local RAS systems, which may partly be independent of systemic RAS (3, 7, 11).

**Figure 1:**
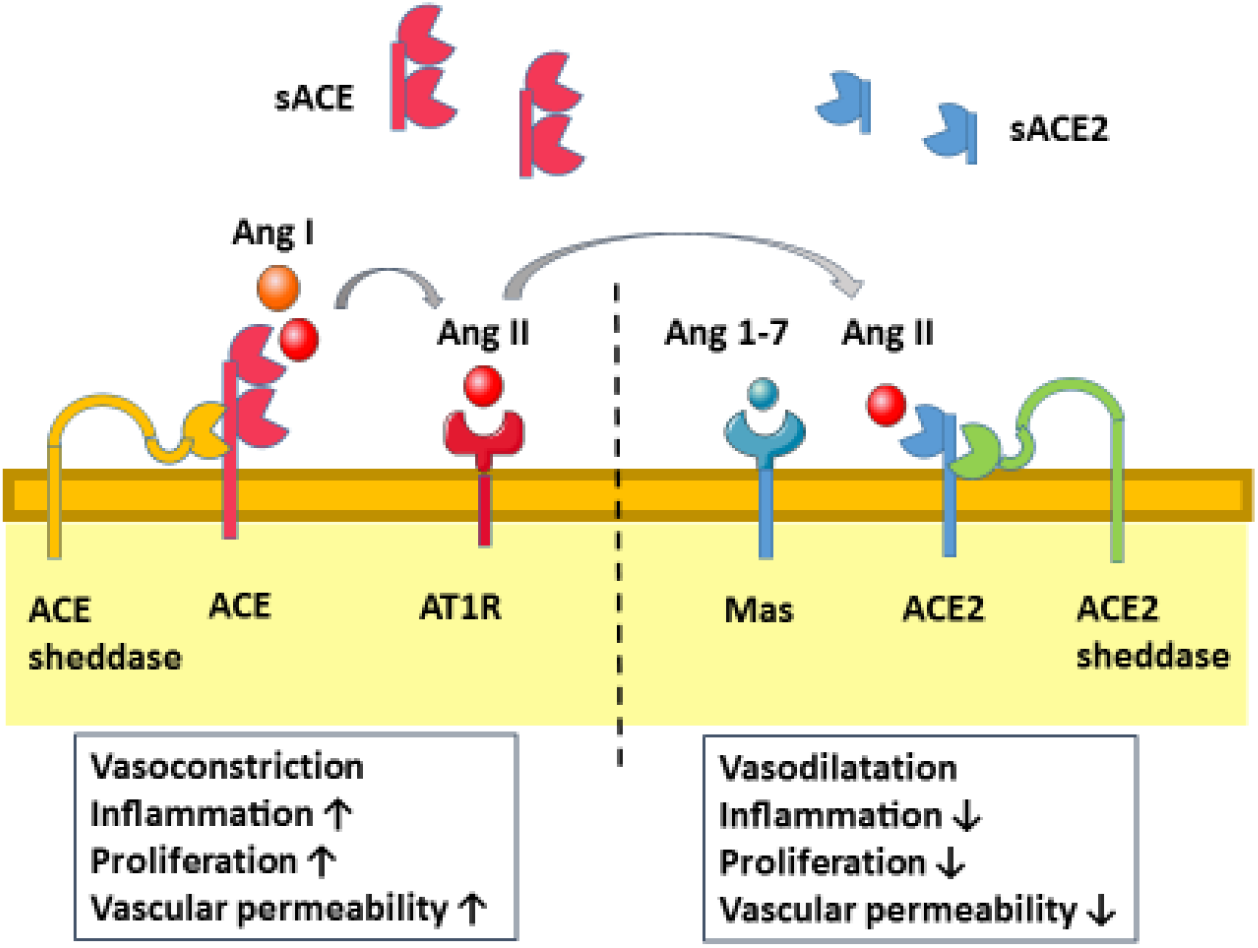
The ACE/Ang II/AT1R arm of the RAS causes vasoconstriction, is prothrombotic and increases inflammation, proliferation and vascular permeability. ACE is shed into plasma primarily from pulmonary microvascular endothelium by an as yet unknown sheddase. Constitutive shedding of sACE is high. The ACE2/Ang 1-7/Mas-receptor arm of RAS counteracts Ang II resulting in vasodilation and reduced inflammation, proliferation and vascular permeability. Cell membrane ACE2 levels are primarily translationally and post-translationally regulated. Constitutive shedding of ACE2 is low and mediated primarily by the sheddase ADAM17 (a disintegrin and metalloproteinase 17). Induced shedding of ACE2 can be caused by different stimuli and may involve other sheddases.

The consequences of SARS-CoV-2 interactions with ACE2 are debated, as are effects on local and systemic RAS balances and how these may contribute to COVID-19 pathology. It has been proposed that 1) high expression of ACE2 may confer increased susceptibility to SARS-CoV-2 and more severe COVID-19 and 2) that a relative lack of ACE2 in infected tissues may exacerbate local effects of Ang II, contributing to increased vasoconstriction, vascular permeability, inflammation and thrombosis (5, 12). Experimental studies of SARS-CoV have shown that virus binding, infection and replication can down-regulate cell surface ACE2 by virus-induced receptor internalization, reduced ACE2 expression and enhanced shedding of ACE2 from the cell membrane (13-16). Similar effects are expected for SARS-CoV-2. Models of severe acute lung injury (ALI) and SARS-CoV have demonstrated a protective role of ACE2, with enhanced lung injury in ACE2 knockouts and under ACE2 inhibition (13, 17). However, a recent autopsy study of human patients who succumbed either to COVID-19 or influenza A (H1N1) unexpectedly showed high ACE2 protein expression in alveolar and pulmonary endothelial cells (18). A possible explanation for this finding could be interferon-induced up-regulation of ACE2 which was found for humans (but not rodents) in transcriptomic studies (19, 20). Notably, many risk factors for severe COVID-19 infection such as male sex, age, hypertension, diabetes and heart failure are associated with chronically elevated levels of soluble ACE2 (21-23), which may reflect enhanced ACE2 shedding and/or increased ACE2 expression in these groups. It is also an unresolved issue whether treatment with RAS-inhibitors influences SARS-CoV-2 susceptibility and/or disease severity (24, 25).

Attention has so far focused on virus-induced changes of ACE2, but signaling in the ACE/Ang II/AT1R arm of RAS is also influenced by pulmonary disease and ALI (11). In animal models of pneumonia, Ang II levels increased in plasma and lungs within hours of injury (17). In human infectious ARDS, ACE activity was substantially increased in bronchoalveolar lavage (BAL) compared with healthy controls (26). Clinical studies of ACE activity in plasma of patients with acute lung injury or ARDS showed a dynamic pattern with decrease over the first days and, for patients who recover, subsequent normalization (27, 28). The degree and duration of the ACE decrease was shown to be associated with disease severity and has been attributed to pulmonary endothelial injury. Models of bacterial pneumonia have demonstrated the importance of a dynamic pulmonary RAS response; ACE2 in BAL and lungs was initially decreased to allow entry of immune cells and then increased to limit vascular permeability and resolve inflammation (29, 30). Both RAS-effector arms may thus be involved in the pathology of COVID-19 pneumonia, and their dynamic temporal responses may influence RAS-balances locally and systemically. Shedding is expected to be a central regulating mechanism for both ACE2 and ACE. The aim of this study was to measure circulating levels of soluble ACE2 (sACE2) and ACE (sACE) during and four months after COVID-19, and to investigate their associations with outcome and risk factors for severe COVID-19. Since both sACE2 and sACE may be released from the inflamed pulmonary endothelium, we also investigated their correlations with markers of inflammation and endothelial dysfunction.

## 2 Methods

### 2.1 Study population

Patients treated for COVID-19 at Danderyd Hospital 15^th^ of April to 11^th^ of June 2020 were offered participation in the COMMUNITY project, as previously described (31-33). Inclusion criteria were COVID-19 diagnosis, either PCR-confirmed (110 patients) or by clinical presentation and typical radiology (4 patients). The only exclusion criteria for the present study was age below 18 years and inability to give informed consent. The study was approved by the National Ethical Board (EPM 2020-01653). Patients were subjected to blood sampling two to three times per week while in hospital; plasma samples were stored in a biobank. A total of 116 patients were recruited with two patients being excluded from the present study: one was also included in an interventional study with transfusion of convalescent plasma and one lacked EDTA plasma samples at first test. Surviving patients were offered a follow-up visit with blood sampling after four months; plasma from 58 patients was obtained in this phase. For comparison ten healthy controls without risk factors for severe COVID-19, RAS-inhibition or cardiovascular disease (CVD) were selected from an existing biobank (approval from local ethical board, EPN Stockholm reference 2015/914-31). Median age of healthy controls was 70.6 (interquartile range IQR 69-72) years and 7/10 were male. All study subjects gave informed consent.

Data on medical history, status, medication, routine laboratory tests, level of care, respiratory support and clinical development were taken from medical records. Death during hospital stay was studied as outcome. Other indicators of disease severity were level of care (normal ward, intermediate care unit, intensive care unit) and level of respiratory support (none, oxygen < 5 l/min, oxygen 5-12 l/min, high flow oxygen or non-invasive ventilation, intubation). Analyzed risk factors for severe COVID-19 infection were age, BMI, male sex, diabetes, hypertension, known CVD and chronic pulmonary disease. The total number of pre-existing comorbidities was individually calculated.

### 2.2 Blood sampling

Blood samples were taken in resting, fasting condition in reclining position in the morning in connection with routine blood sampling. They were centrifuged at 2000 g for 20 minutes at room temperature within two hours of blood sampling and immediately stored at −80^°^C until analyses.

### 2.3 Soluble ACE2 and ACE measurements

Concentrations of soluble ACE2 and ACE were measured by commercially available sandwich enzyme-linked immunosorbent assays (ELISAs): ACE2 human ELISA kit (Adipogen Life Sciences, Switzerland, catalog number AG-45B-0023-KI01) and human ACE ELISA kit (XpressBio, MD, USA, catalog number XPEH0026). EDTA plasma aliquots were thawed on ice and analyzed in duplicate according to the manufacturer’s instructions in a biosafety hood level two. For ACE2 analyses, patient plasma was diluted 1:4 and plasma from healthy controls 1:2. For ACE analyses all samples were diluted 1:8. Optical density was read by a Tecan platelet reader and concentrations were calculated by interpolation from the standard curve. Samples with an OD above the highest point in the standard curve were linearly extrapolated up to the maximum standard concentration plus 50%, else set to this value.

### 2.4 Markers of inflammation and endothelial activation

Routine laboratory tests included CRP, blood counts, creatinine, sodium and potassium. For subsets of patients, procalcitonin and leukocyte differential counts were available as routine tests taken at the same blood sampling occasion. Von Willebrand factor (VWF), fibrin D-dimer, factor VIII (fVIII) and plasminogen activator inhibitor 1 (PAI-1) were analyzed in another study of the COMMUNITY project, as has been reported (31). The same study also quantified interleukin 6 (IL-6) and tumor necrosis factor α (TNFα) using commercially available ELISAs from R&D systems (Bio-techne, Minneapolis, MN, USA).

### 2.5 Statistical analyses

sACE2 and sACE concentrations are presented as median with IQR as both were positively skewed. Other continuous variables are presented as mean ± standard deviation if normally distributed, otherwise as median with IQR. Categorical variables are presented as numbers and proportions. Differences between independent groups were analyzed by Mann-Whitney U-test. Differences between sACE2 and sACE at recruitment compared to at follow-up were analyzed by Wilcoxon signed rank test. Correlations between continuous variables were analyzed by Spearman’s correlation coefficient. Differences in sACE2 respectively sACE depending on categorical variables were analyzed univariately by Mann-Whitney U-test or Kruskal-Wallis test, as appropriate. P-values < 0.05 were considered significant. Statistical analyses were performed in SPSS version 26 (IBM, United States) and figure 2 generated by Prism (Graphpad Software Inc., California, United States).

**Figure 2:**
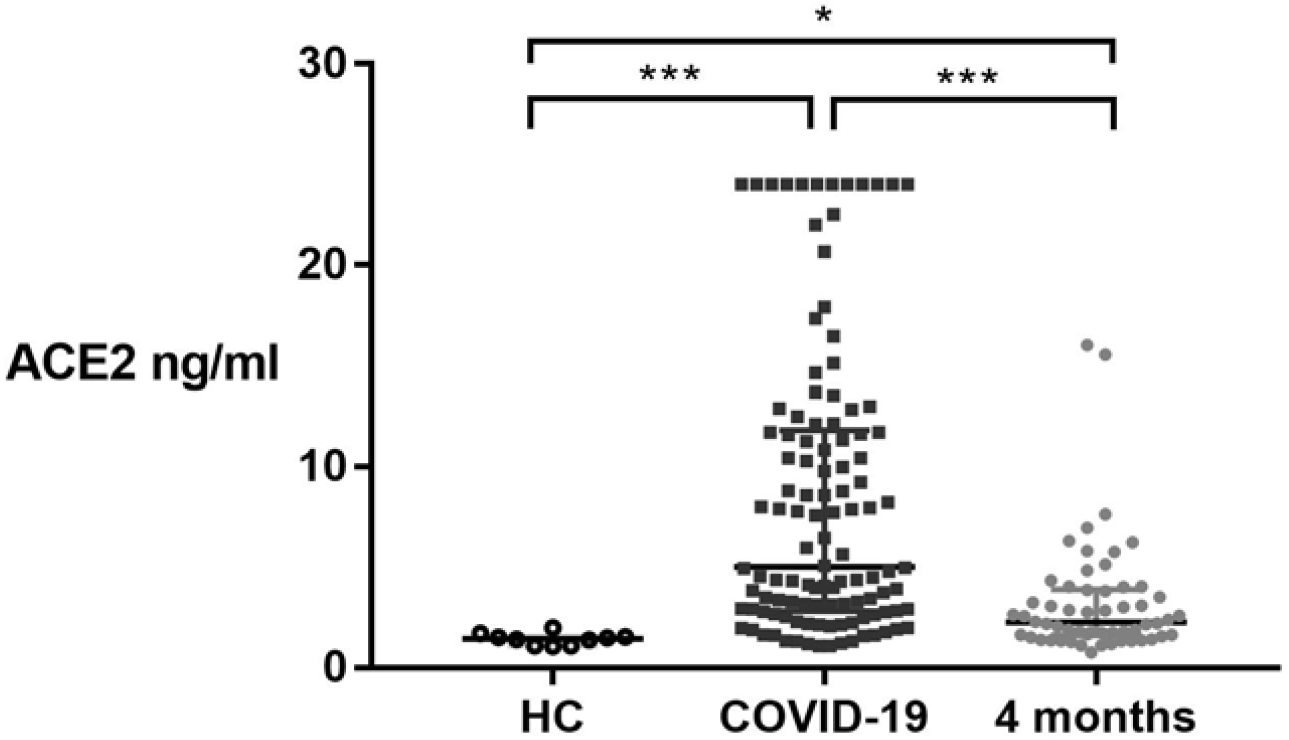
sACE2 concentrations for 10 healthy controls (HC, left), 114 patients with COVID-19 (middle) and 58 patients at follow-up after four months (right). The differences between COVID-19 patients and HC were analyzed by Mann-Whitney U-test. The difference between acute COVID-19 and follow-up four months later was analyzed by Wilcoxon signed rank tests. ***) p < 0.0001, *) p < 0.05.

## 3 Results

### 3.1 Cohort characteristics

The characteristics of the 114 COVID-19 patients sampled at recruitment are shown in table 1. The majority (64%) were male and mean age was 59 years. The most common symptoms at admittance were cough (75%), dyspnea (75%) and fever (68%); diarrhea (28%) and myalgia (28%) were less common. Mean symptom duration at first blood sampling was 11.5 ± 6 days. Patients had median one pre-existing comorbidity (IQR 0-2), the most common being diabetes, hypertension and cardiovascular disease. Asthma was the most common chronic pulmonary disease. Twenty-five patients (22%) were treated at the intermediate or intensive care unit during their hospital stay. Twelve patients (10.5%) were intubated and thirteen (11.4%) patients died, twelve men and one woman.

**Table 1:**
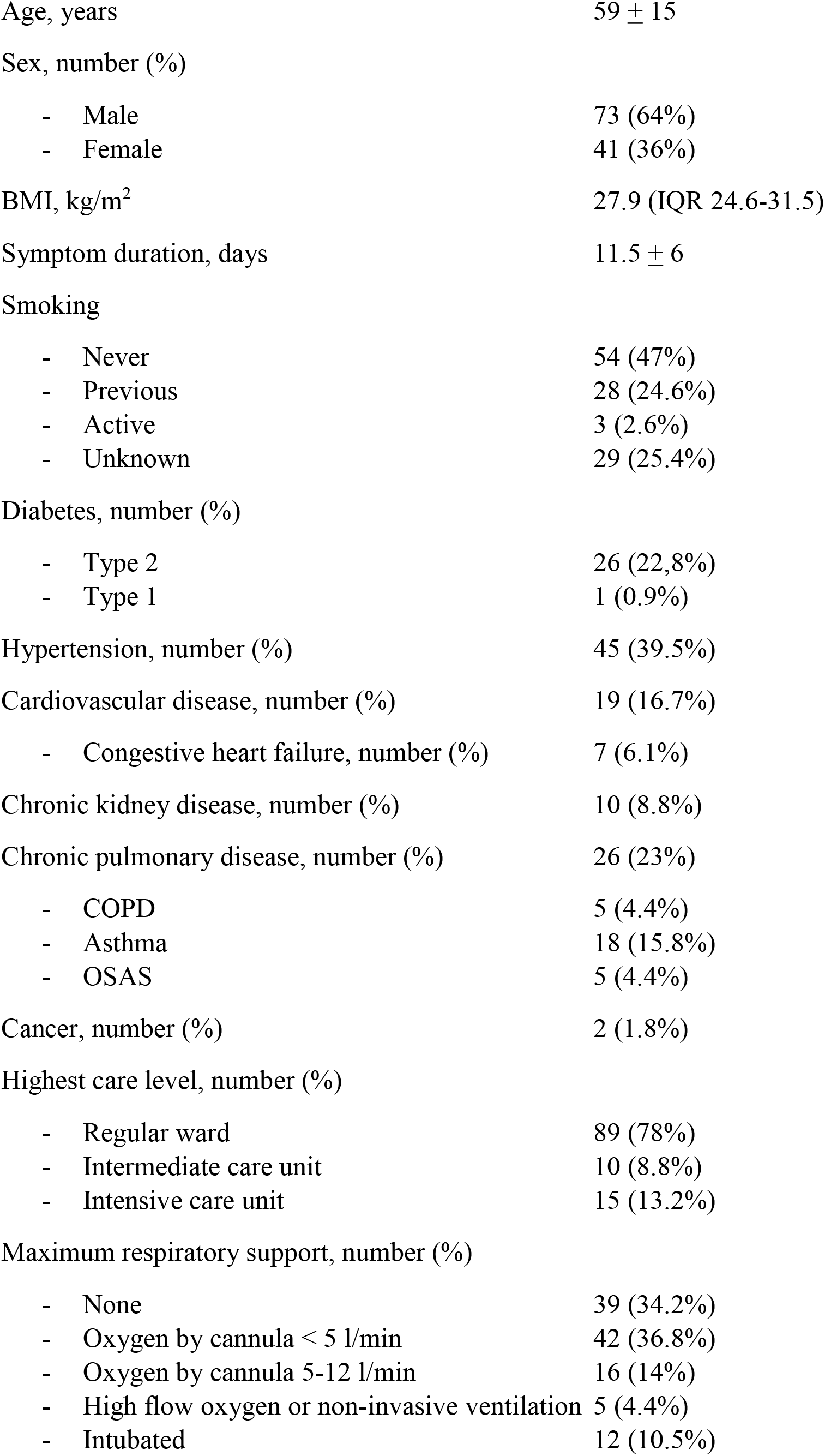
Cohort characteristics (N=114) presented as mean ± standard deviation, median (interquartile range) or number (%). BMI: body mass index; COPD: chronic obstructive pulmonary disease; OSAS: obstructive sleep apnea syndrome.

|  |  |
| --- | --- |
| Age, years | 59 ± 15 |
| Sex, number (%) |  |
| - Male | 73 (64%) |
| - Female | 41 (36%) |
| BMI, kg/m<sup>2</sup> | 27.9 (IQR 24.6-31.5) |
| Symptom duration, days | 11.5 ± 6 |
| Smoking |  |
| - Never | 54 (47%) |
| - Previous | 28 (24.6%) |
| - Active | 3 (2.6%) |
| - Unknown | 29 (25.4%) |
| Diabetes, number (%) |  |
| - Type 2 | 26 (22.8%) |
| - Type 1 | 1 (0.9%) |
| Hypertension, number (%) | 45 (39.5%) |
| Cardiovascular disease, number (%) | 19 (16.7%) |
| - Congestive heart failure, number (%) | 7 (6.1%) |
| Chronic kidney disease, number (%) | 10 (8.8%) |
| Chronic pulmonary disease, number (%) | 26 (23%) |
| - COPD | 5 (4.4%) |
| - Asthma | 18 (15.8%) |
| - OSAS | 5 (4.4%) |
| Cancer, number (%) | 2 (1.8%) |
| Highest care level, number (%) |  |
| - Regular ward | 89 (78%) |
| - Intermediate care unit | 10 (8.8%) |
| - Intensive care unit | 15 (13.2%) |
| Maximum respiratory support, number (%) |  |
| - None | 39 (34.2%) |
| - Oxygen by cannula < 5 l/min | 42 (36.8%) |
| - Oxygen by cannula 5-12 l/min | 16 (14%) |
| - High flow oxygen or non-invasive ventilation | 5 (4.4%) |
| - Intubated | 12 (10.5%) |

### 3.2 Soluble ACE2

Levels of sACE2 were higher in patients than healthy controls, median 5.0 (IQR 2.8-11.8) ng/ml versus 1.4 (1.1-1.6) ng/ml, p < 0.0001, see figure 2. There was a positive correlation between sACE2 levels and symptom duration at testing r=0.35, p < 0.0001. sACE2 concentrations were significantly higher for men than women, median 7.7 versus 3.8 ng/ml, p=0.021. Other risk factors for severe COVID-19 infection did not result in higher sACE2 levels by univariate analysis, see table 2. There was no correlation between sACE2 and total number of comorbidities. sACE2 levels were not affected by treatment with RAS-inhibitors (n=37), neither for ACE inhibitors (ACEI, n=19) nor AT1R receptor blockers (ARB, n=18), data not shown. There were no correlations between sACE2 and vital parameters (heart rate, respiratory frequency, blood pressure) at the time of blood sampling (data not shown). Levels of sACE2 did not differ significantly between patients who died and survivors, and also did not depend on other indicators of COVID-19 severity (care level or required respiratory support, data not shown).

**Table 2:** Univariate analysis of acute sACE2 (n=114) respectively sACE2 at follow-up (n=58) after four months (sACE follow-up) versus risk factors for severe Covid-19. (N): number of patients with presence of the risk factor. Continuous variables: r= Spearman’s correlation coefficient. Categorical variables: percentage difference in median sACE2 concentration in presence of the risk factor, p-value calculated by Mann-Whitney U-test. BMI: body mass index, CVD: cardiovascular disease. NS: not significant, NA: not applicable.

|  | sACE2<br>N=114 | p-value | sACE2<br>follow-up<br>N=58 | p-value |
| --- | --- | --- | --- | --- |
| Age | r= -0.025 | 0.792, NS | r=0.288 | 0.029* |
| BMI | r= 0.020 | 0.838, NS | r=0.347 | 0.009** |
| Total no of comorbidities | r=0.118 | 0.212, NS | r=0.399 | 0.002** |
| Male sex (N=73/114; 40/58 ) | + 102% | 0.021* | NA | 0.181, NS |
| Hypertension (N=44/114; 21/58) | NA | 0.974, NS | + 61% | 0.062 |
| Diabetes (N=27/114, 11/58) | NA | 0.826, NS | + 86% | 0.048* |
| CVD (N=19/114, 9/58) | NA | 0.153, NS | NA | 0.225, NS |
| Chronic pulm dis (N=26/114, 11/58) | NA | 0.230, NS | NA | 0.433, NS |
| Smoking (N=3/114, 2/58) | NA | 0.237, NS | NA | 0.558, NS |

At follow-up, mean four months (121 ±14 days) after initial bloodsampl, median sACE2 decreased to 2.3 (1.6-3.9) ng/ml (p < 0.0001) for the 58 patients sampled at both occasions, but remained significantly higher in patients than in healthy controls (p=0.012), see figure 2. sACE2 at recruitment and sACE2 at follow-up were moderately correlated, r=0.44, p < 0.0001. By univariate analysis, follow-up sACE2 correlated significantly with age and BMI and was higher for patients with diabetes than those without, see table 2. Follow-up sACE2 was higher for patients with RAS-inhibitors than those without, 3.1 (2.4-4.9) versus 1.9 (1.4-3.3) ng/ml, p=0.028 (n=16/58) and correlated with total number of comorbidities, see table 2.

### 3.3 Soluble ACE

Levels of sACE in COVID-19 patients were not significantly different from healthy controls, 57 (45-70) ng/ml versus 64 (48-265) ng/ml. There was no difference in sACE levels depending on risk factors for severe COVID-19 by univariate analysis (data not shown). There was a weak correlation between sACE and total number of comorbidities, r=0.21, p=0.023. Levels of sACE were not affected by treatment with RAS inhibitors and did not correlate with vital parameters (data not shown). Levels of sACE did not differ depending on survival or other indicators of disease severity.

sACE at follow-up increased marginally to 72 (52-87) ng/ml, p=0.008 (n=58). There was a weak to moderate correlation between sACE at recruitment and sACE at follow-up, r=0.377, p=0.004. Levels of follow-up sACE did not differ depending on risk factors for severe COVID-19 infection and did not correlate with total number of comorbidities. Also, follow-up sACE did not differ depending on treatment with RAS-inhibitors.

There was no significant correlation between sACE2 and sACE during COVID-19 (p=0.788), but a trend to weak positive correlation between sACE2 and sACE at follow-up, r=0.25, p= 0.06.

### 3.4 Correlations of sACE2 and sACE versus markers of inflammation and endothelial dysfunction

Spearman’s correlation coefficients were calculated for sACE2 and sACE versus markers of endothelial activation and inflammation. Significant correlations with │r│> 0.25 are presented in table 3. Neither sACE2 nor sACE correlated significantly with CRP (n=110), procalcitonin (n=67), neutrophil count (n=98) or lymphocyte count (n=97). Only sACE2 had a weak positive correlation with white blood cell count. ACE2 correlated positively with monocyte and platelet counts. In contrast, acute sACE displayed negative correlations with monocyte and platelet counts. There were positive correlations between sACE2 and V WF, fVIII and D-dimer that were absent for sACE. sACE instead displayed positive correlations with IL6, TNFα and PAI-1 which were not seen for sACE2.

**Table 3:** Spearman correlation coefficients for significant correlations between acute sACE2 respectively sACE and markers of inflammation and endothelial dysfunction. Correlation coefficients with│r│> 0.25 are reported. N: number of patients analyzed. WBC: white blood cell count; VWF: von Willebrand factor; PAI-1: plasminogen activator inhibitor 1; IL-6: interleukin 6; TNFα: tumor necrosis factor α.*) p 0.01-0.049, **) p 0.001-0.0099, ***) p < 0.001.

|  | ACE2 | ACE |
| --- | --- | --- |
| WBC (n=109) | 0.26** | NS |
| Monocytes (n=97) | 0.39*** | - 0.26* |
| Platelet count (n=109) | 0.32** | - 0.34** |
| VWF (n=110) | 0.36*** | NS |
| Factor VIII (n=110) | 0.55*** | NS |
| D-dimer (n=110) | 0.30** | NS |
| IL-6 (n=99) | NS | 0.43*** |
| TNFα (n=100) | NS | 0.43*** |
| PAI-1 (n=103) | NS | 0.44*** |

## 4 Discussion

The main finding of this study was that levels of sACE2 were strongly elevated in COVID-19 patients but approached normal levels four months after infection. sACE2 during admission was higher in men than women but was not affected by other risk factors for severe COVID - 19. In contrast, sACE2 levels at follow-up were higher in relation to several risk factors and under treatment with RAS-inhibitors, and also correlated with total number of comorbidities. sACE2 and sACE levels did not differ depending on COVID-19 outcome or disease severity. sACE2 and sACE differed distinctly in their correlations with markers of inflammation and endothelial dysfunction, which may imply release from different cell types.

To our knowledge, sACE2 antigen has not been previously measured in the acute phase of COVID-19. A recent study showed plasma ACE2 activity to be strongly elevated one month after COVID-19 compared with controls (34). In contrast with our results, levels were maintained for several months in patients who were followed up repeatedly. sACE2 maintains enzymatic activity in airway surface liquid and cell culture media (16), but in plasma sACE2 activity is limited by an as yet unknown endogenous ACE2 inhibitor (35). The method of Patel et al is based on pre-analytical removal of the endogenous ACE2 inhibitor and may not reflect in vivo activity. It thus remains an open question how circulating ACE2 levels in COVID-19 influence circulating Ang II and Ang 1-7 levels. An early study of 12 COVID-19 patients reported high Ang II levels which correlated with viral load and disease severity (36), while another study found no difference in Ang II between COVID-19 patients and healthy controls (37). Measurement of Ang II is technically demanding due to short half-life which may contribute to the discrepancy. Henry et al found low concentrations of angiotensin I and Ang 1-7 in COVID-19 patients compared with healthy controls (38). Combined, the above studies indicate that high circulating sACE2 does not overcome the effects of membrane-based ACE2 and/or ACE during COVID-19, but further research is needed.

Shedding was the proposed mechanism behind high plasma ACE2 activity after COVID-19 (34), and virus-induced shedding of ACE2 has previously been demonstrated in several studies of SARS-CoV (14-16, 39). Shedding of ACE2 in airway epithelial cells occurs constitutively but is also inducible (6, 14, 16, 40). Different stimuli increase ACE2 shedding, including cytokines and hypoxia (6, 16, 41, 42). Ang II also increases ACE2 shedding (43) and potentially high levels of Ang II in infected tissues can cause shedding of ACE2 which may reach the circulation due to increased vascular permeability. ADAM17 (A disintegrin and metalloproteinase 17) is considered to be the main sheddase for ACE2 (39); other sheddases may also contribute to inducible shedding (16). It has been proposed that ADAM17 activation in infected cells could be a major factor in COVID-19 pathology, potentially contributing to the cytokine storm (44, 45). We found no significant correlation between acute ACE2 and TNFα, one of the main substrates of ADAM17, providing no support for this hypothesis. It remains to be determined which cell types and shedding mechanisms contribute release of ACE2 into the circulation in COVID-19 but pulmonary endothelial and/or alveolar cells are likely sources. Recent finding suggest that not only shedding, but also increased ACE2 transcription and expression could contribute to high sACE2 levels in COVID-19. Ackerman et al found high ACE2 protein levels in pulmonary endothelial and alveolar cells in deceased COVID-19 and influenza H1N1 patients (18). Several studies have also demonstrated interferon-induced increase of ACE2 transcription in human epithelial cells (20) suggesting that membrane-based ACE2, contrary to what has been assumed, may be increased in COVID-19.

Unexpectedly sACE2 levels during COVID-19 did not differ depending on the presence of risk factors for severe COVID-19 infection (with the exception of male sex) and were not affected by RAS inhibition. In contrast, sACE2 four months later correlated with age and BMI and was higher for patients with diabetes, as found by others (21-23). Moreover, sACE2 at follow-up was higher under treatment with RAS-inhibitors and correlated with the total number of comorbidities. COVID-19 induced release of sACE2 thus appears non-discriminate, erasing associations between sACE and most risk factors for severe COVID-19. Male sex deviated from this pattern, with clearly higher sACE2 levels in men than in women. The study by Patel et al and two studies of heart failure also found higher circulating sACE2 antigen/activity in men as compared to in women (34, 46, 47) suggesting that ACE2 shedding may be higher in men under pathological conditions. It warrants further investigation if increased shedding or high sACE2 levels contribute to more severe disease in men. In our study sACE2 levels did not differ depending on survival or other indicators of disease severity. Groups with severe or fatal COVID-19 were small and power may have been insufficient to detect differences. However, the result may also reflect the dual nature of ACE2 in COVID-19 (5). High sACE2 levels may reflect both a high viral load and sufficient remaining tissue ACE2 to resolve inflammation. Furthermore, sACE2 reduces infectivity of both Sars-CoV (16, 48) and Sars-CoV-2 (49), suggesting that high sACE2 levels could limit secondary viral establishment.

Differences in sACE during COVID-19 were marginal, with slightly lower sACE during infection compared with four months later. In previous studies of ALI and ARDS, more pronounced decreases of plasma ACE activity were seen, which were attributed to injury of pulmonary microvascular endothelial cells (27, 28). The observed decrease could thus be non-specific.

Correlations between sACE2 respectively sACE and markers of inflammation and endothelial dysfunction were investigated due to the inflammatory effect of Ang II, the anti-inflammatory effect of Ang 1-7, ACE’s role in the immune system (50) and the fact that the inflamed (pulmonary) endothelium may shed both sACE2 and sACE. Correlations differed distinctly for sACE2 and sACE. sACE2 and sACE had opposite correlations with monocytes and platelets, with sACE2 being positively and sACE negatively correlated with both. We speculate that processes resulting in shedding of ACE2 respectively ACE could be associated with different endothelial adhesive properties, monocyte transmigration and/or vascular permeability. Concerning platelets, recent studies have demonstrated ACE2 expression by platelets, with a possibility for SARS-CoV-2 platelet infection which could theoretically result in ACE2 shedding (51, 52). sACE2 displayed positive correlations with VWF, fVIII and D-dimer, which were absent for sACE. VWF is thought to be released from endothelial activation in COVID-19, and the results imply that release of sACE2 to a greater extent than sACE depends on endothelial activation. In contrast sACE correlated positively with IL-6, TNFα and PAI-1, which sACE2 did not. Possibly ACE release involves different type of endothelial injury or release form other vascular beds than sACE2. sACE is also expressed in immune cells, in particular monocytes and macrophages, which could thus be an alternative sources of sACE. It warrants further investigation if COVID-19 causes if a disturbed RAS-balance in the immune system and if this contributes to hyper-inflammation and pathology.

### Limitations

The study is of moderate size and included hospital-treated patients with a limited proportion severe/fatal COVID-19. While the cohort is considered representative, it was heterogenous with respect to age and underlying conditions, which makes it potentially vulnerable to confounding. There are no standardized methods to measure sACE2 antigen. ELISA results were stable with median intra-assay CoV 1.8% (0.8-3.1%) between wells and sACE2 values were similar to those of a previous study (53). We did not have spare aliquots to rerun tests at higher dilution for patients with OD above maximum concentration on the standard curve; concentrations were therefore likely underestimated for ACE2 in 16 patients and for ACE in 10 patients. The group of healthy controls could have been larger considering that levels of sACE depend on genetic polymorphisms. However, our main conclusions are based on comparisons of values during COVID-19 compared with four months later with patients being their own controls.

In conclusion, we find the plasma RAS-balance in hospital-treated Covid-19 patients to be characterized by a strong transient increase of circulation plasma sACE2 combined with marginally reduced sACE. Contributing factors to the sACE2 elevation likely include increased shedding in infected cells, and possibly also increased ACE2 membrane expression. The reduction of sACE may not be specific for COVID-19 and could be secondary to pneumonia. The relation between circulation and tissue ACE2 needs further investigation, but can be assumed to involve a relative lack of tissue ACE2. Effects on angiotensin peptides as well as metabolism of bradykinin and its metabolites are difficult to predict and require further study. Treatment with RAS-inhibitors had no significant effect on sACE2 levels during COVID-19 and future studies on interventions targeting the RAS-imbalance may consider means to limit or compensate for ACE2 shedding, for instance by ADAM17-inhibition or substitution Ang 1-7.

## Data Availability

Data may be made available upon the completion of the COMMUNITY study.

## Acknowledgements

We are grateful for the assistance of BMA Martha Kihlgren for sACE2 and sACE analyses. Blood sampling and plasma preparation by research nurses Lena Gabrielsson, Nina Greilert, Eva Isaksson and BMA Ann-Christin Salomonsson is gratefully acknowledged.

